# Risk of adverse pregnancy outcomes and impact of statin use in pregnant women with familial hypercholesterolemia

**DOI:** 10.1101/2024.09.03.24312275

**Authors:** Karianne Svendsen, Jacob Juel Christensen, Jannicke Igland, Henriette Walaas Krogh, Liv J. Mundal, David R. Jacobs, Martin P. Bogsrud, Kirsten B. Holven, Kjetil Retterstøl

**Affiliations:** Lipid Clinic, Oslo University Hospital, Oslo, Norway; Department of Nutrition, Institute of Basic Medical Sciences, University of Oslo, Oslo, Norway; Cancer Registry of Norway, Norwegian Institute of Public Health, 0379 Oslo, Norway; Department of Global Public Health and Primary Care, University of Bergen, Bergen, Norway; Department of Health and Social Science, Centre for Evidence-Based Practice, Western Norway; Department of Health and Caring Sciences, Western Norway University of Applied Sciences, Bergen, Norway; Department of Non-Communicable Diseases, Division of Prevention and Public Health, Norwegian Directorate of Health, Oslo, Norway; University of Minnesota, Minneapolis, Minnesota, USA; Unit for Cardiac and Cardiovascular Genetics, Oslo University Hospital, Oslo, Norway; Norwegian National Advisory Unit on Familial Hypercholesterolemia, Oslo University Hospital, Oslo, Norway

**Keywords:** Familial hypercholesterolemia, adverse pregnancy outcomes, statins, low birthweight, premature births, preeclampsia, diabetes, bleedings, c-section, malformations

## Abstract

**Background and aims:** Sparse data exist on the possible risk of adverse pregnancy outcomes in women with familial hypercholesterolemia (FH). We investigated associations between having a FH diagnosis and adverse pregnancy outcomes, and between statin exposure in pregnancy and adverse pregnancy outcomes among women with FH.

**Methods:** This registry-based study included 3869 pregnancies among 1869 women with FH and 68225 pregnancies among 33661 women from the general population. Data on adverse pregnancy outcomes were obtained from the Medical Birth Registry of Norway with data from 1967-2018. Data on pharmacy-dispensed statins were obtained from the Norwegian prescription database (2004-2018) in 1051 women with FH. Associations were presented as odds ratio (OR) with 95% CI from logistic regression adjusted for mother’s age, parity, and offspring’s birth year.

**Results:** Women with FH had a higher risk of preeclampsia (OR 1.21 [1.00-1.46]), but lower risk for gestational diabetes (OR 0.58 [0.36-0.92]) and intrapartum hemorrhage during delivery (OR 0.81 [0.71-0.92]) compared to controls. No excess risk of adverse pregnancy outcome in offspring was observed for FH vs controls. Women with FH using statins in pregnancy (n=260) had a higher risk of having offspring with low (<2500 g) birth weight (OR 2.20 [1.11, 4.49]) compared to non-exposed women with FH (n=791).

**Conclusions:** Women with FH had lower risk of gestational diabetes and intrapartum hemorrhage during delivery and non-significantly higher risk of preeclampsia compared to controls. No difference in adverse pregnancy outcomes in the offspring was observed. Statin exposure in pregnancy was associated with a higher risk of having offspring with low birth weight among women with FH, and this association warrants further investigations.

## Introduction

Individuals with familial hypercholesterolemia (FH) have an inherited condition that reduces the capacity to clear LDL-cholesterol (LDL-C) particles from the circulation ^1^, leading to accelerated atherosclerosis, and excess risk of premature coronary heart disease (CHD) ^2, 3^. There is a physiological increase in blood lipids during pregnancy, and statins are contraindicated during pregnancy and lactation ^4^. Thus, women with FH may experience significantly heightened levels of LDL-C without the option of statin treatment ^5, 6^. The potential atherogenic consequences of this dual increase in LDL-C in women with FH are incompletely understood ^7^.

Although hypercholesterolemia is not normally mentioned as a risk factor ^4^, it has been linked to higher risk of preeclampsia ^8, 9^, and blood lipid dysregulation is again strongly associated with gestational diabetes ^7^. Since diet and lifestyle are significant predictors of both hypercholesterolemia, preeclampsia and gestational diabetes ^10, 11^, genetically verified FH can serve as a model for studying the impact of hypercholesterolemia in isolation. Using Norwegian health registry linkage in a sub-sample of the present FH population, it was reported that offspring from mothers with FH who carried to full-term did not have a higher risk of selected adverse pregnancy outcomes ^12^. However, diagnostics of these conditions have improved over time, and it is unknown whether the risk of preeclampsia, gestational diabetes and gestational hypertension differ between women with and without genetically verified FH today.

Due to the limited information on safety of statin use during pregnancy and lactation, a precautionary and individualized approach comparing risks and benefits of statin use in pregnancy and during lactation is recommended ^4, 13, 14^. Because many women may not realize they are pregnant until a few weeks after conception, some women might unknowingly have exposed their offspring to statins. This is mostly relevant in women with FH who is recommended to start lifelong statin therapy in adolescence ^15, 16^. About 30% of a subsample of Norwegian women with FH have used statins in their early pregnancy ^17^. This represents a challenge for statin treated women who worry about potential adverse effects of statin use on their offspring ^17–19^. Although some studies have shown no teratogenic effect of maternal statin use ^18, 20^, other studies of women from the general population have suggested that statin use may be associated with premature birth and low birth weight ^21, 22^. Hence, in terms of the association between FH and adverse pregnancy outcomes and statin use in pregnancy, there is still little robust data to ease the worry.

The aim of this study was to compare women with FH and controls on terms of risk of adverse pregnancy outcomes for mothers and offspring, and to investigate associations between statin exposure in pregnancy and adverse outcomes among women with FH.

## Methods

### Study design and variables

The study originates from a registry-based matched cohort-study of individuals with a genetically verified FH diagnosis from the Unit for Cardiac and Cardiovascular Genetics (UCCG) database at Oslo University Hospital ^23^. The controls (1:20) were drawn from the general Norwegian population matched on sex and year of birth ^24^. Women from these cohorts with one or more pregnancies were included in the present study.

Data on pregnancy characteristics and adverse pregnancy outcomes during 1967-2018 were obtained from The Medical Birth Registry (MBRN) ^25^. This registry contains data on women’s health during pregnancy, childbirth and in the maternity period, and children’s health from birth and up to one year of age ^25^. In the present study, for all available pregnancies, we obtained from the MBRN information on mother’s birth year, age at birth, height and weight before-and at end of pregnancy (data available from 2007 and only from about 40% of the hospitals), smoking habits (data available form 1999 but with <50% of the data missing), mother’s health prior to pregnancy (including occurrence of diabetes mellitus [type 1, 2 or unspecified] and chronic hypertension), previous spontaneous abortions (before week 12 and 12-23), and last menstruation period. Data on pregnancy characteristics such as date of birth and age of mothers, body mass index (BMI) at start and end of pregnancy (BMI information was only available from 2007 and from about 40% of all institutions), in vitro fertilization (IVF), preeclampsia, gestational hypertension, gestational diabetes, intrapartum hemorrhage (defined as ≥ large bleedings 500 ml during delivery) and delivery methods were also included. Furthermore, the offspring’s date of birth, birth weight, gestational length (days and weeks), preterm births (defined as births prior to gestational week 37), mean birthweight and different categories of birth weight (low birth weight (<2500 grams), high birth weight (>4500 grams), birthweight <10^th^ and 25^th^ percentiles), Appearance, Pulse, Grimace, Activity, and Respiration (APGAR) scores (tests of newborns immediately after delivery), complications during birth, occurrence of various malformations combined to one group “any malformation”, congenital heart defects, stillbirths and neonatal deaths.

The Norwegian Prescription database contains information on pharmacy-dispensed prescriptions ^26^. We obtained data on dispensing date, year, number of packages, number of daily defined doses, category, category numbers, reimbursement code, package size, drug strength and daily defined doses of the Anatomical Therapeutic Chemical (ATC) codes C10AA and C10BA (statins and statins in combination with ezetimibe) during 2004-2018. Statin use before, during and after pregnancy was estimated by linking information on statin use with data from the MBRN on last menstruation period, birth date and gestational length. Statin use during pregnancy was defined as filling at least one prescription for statins either during pregnancy or before conception if the prescription package duration would last into pregnancy, assuming one tablet per day (here termed overlap).

### Study population

There were 4112 pregnancies among 1926 women with FH and 72727 pregnancies among 34389 controls. The exclusion criteria were pregnancies with missing gestational length or gestational length <22 weeks, as well as those with missing birth weight and plural pregnancies. The final dataset available for analysis comprised 3869 pregnancies among 1869 women with FH, and 68225 pregnancies among 33661 women in the control group (Figure 1).

In the analysis of statin exposure, only those pregnancies with birth year between 1.1.2005-31.12.2018 were included to have complete coverage on prescription data up to one year prior to birth. In total, 1051 pregnancies among women with FH were available for these analyses.

### Statistical analysis

Pregnancy characteristics were presented with mean and standard deviation (SD) or frequency and percentages (%). Associations between FH status and adverse pregnancy outcomes (all binary variables), and statin exposure and adverse pregnancy outcomes were analyzed with logistic regression and presented as odds ratio (OR) with 95% confidence intervals (95% CI). Confidence intervals were estimated using clustered robust standard errors to take into account dependencies between pregnancies for women with more than one pregnancy. CI overlapping 1 indicated non-significance, corresponding to a significance level equal to 5%. Data were analyzed unadjusted and adjusted for the mothers’ age at pregnancy, parity and the children’s birth year (in 5-year categories), and results from the adjusted models were reported as the main results. Using the same method as described above, we did a sensitivity analysis of the interaction between adverse birth effects and known FH status.

Data management and analyses were performed in STATA version 18 (StataCorp. 2023. Stata Statistical Software: Release 18. College Station, TX: StataCorp LLC.) and R Studio version 2023.03.1 (Posit PBC, Boston, MA, USA).

### Consent

The implementation of the study complies with the Declaration of Helsinki. Consent was required to be included in the UCCG database, and no additional consent was needed to be included in the present study. However, individuals in the FH cohort had the opportunity to withdraw from the study until May 1, 2014. Consent was not needed to be included in the control cohort.

## Results

### Pregnancy-related characteristics

The age at genetically confirmed FH diagnosis was mean 38.9 (14.9) years. At time of delivery, women with FH were on average 27.8 (5.4) years and controls 27.9 (5.4) years. Parity, reported BMI at the start and end of pregnancy, number of smokers and previous spontaneous abortions were similar among women with and without FH. In total 0.5% (n = 21) of women with FH had chronic hypertension, and 0.1% (n = 5) diabetes mellitus before pregnancy, whereas the corresponding percentages in controls were 0.3% (n = 238) and 0.4% (n= 291) (Table 1). The proportion with in vitro fertilization was 0.8% (n = 30) in women with FH and 1.1% (n = 749) in controls (p=0.06).

### Adverse pregnancy outcomes

A higher proportion of women with FH (3.4% n = 133) than controls (2.9%, n = 1951) were diagnosed with preeclampsia during pregnancy (OR 1.21 [95% CI 1.00-1.46]). Gestational diabetes was observed in 0.6% (n = 22) of pregnancies in women with FH, and in 1% (n = 679) of pregnancies in controls (OR 0.58 [95% CI 0.36-0.92]). A lower proportion of women with FH (8.5%, n= 328) experienced intrapartum hemorrhage during delivery compared to controls (10.0 %, n = 6876) (OR 0.81 [95% CI 0.71-0.92]). The frequency of gestational hypertension and cesarean deliveries (c-sections) were similar among cases and controls (Figure 2).

Mean birth weight, proportion with low-and high birth weight, and preterm birth were all similar in offspring from mothers with FH and controls. Similarly, there were no differences in any malformations, stillbirths, or neonatal deaths between offspring of women with and without FH (Table 1 and Figure 3).

APGAR scores were similar between FH and controls: APGAR 1 was 8.7 (SD 1.2) in both FH and controls, and APGAR 5 was 9.3 (SD 0.9) in FH and 8.7 (SD 1.2) in controls.

### Sensitivity analysis

FH was diagnosed prior to pregnancy in 27.6% (n=1067) of all pregnancies of which the majority (n=867) were diagnosed in 2005 or earlier (the earliest available date for diagnosis was 1992). For 34 women, FH was diagnosed during pregnancy.

When stratifying the analyses by pregnancies with births before or equal to 2005 (n = 2818 in FH and 50184 in controls) and after 2005 (n = 1051 in FH and 18040 in controls), as a proxy for unknown versus known FH during pregnancy, the ORs for adverse pregnancy outcomes were like those of the total study population. E.g., the ORs for preeclampsia were 1.24 (95% CI: 0.99-1.54) and 1.14 (95% CI: 0.80, 1.61) and the ORs for gestational diabetes were 0.58 (95% CI: 0.24, 1.42) and 0.58 (95% CI: 0.34, 0.99) between cases and controls with offspring born before and after 2005, respectively.

### Statin use in pregnancy in women with FH

Of the 1051 pregnancies during 2005-18 among women with FH, 37.4% (n = 393) were exposed to statins 6-12 months prior to conception, and 25% (n = 260) during pregnancy (Table 2). The frequency of dispersion of statin prescriptions in all women with FH from one year before conception to one year after birth is visualized in Figure 4.

Statin-exposed (n = 260) and non-exposed (n = 791) pregnancies in women with FH had similar mean age at birth and mean BMI at end of pregnancy, whereas mean BMI at the start of pregnancy was slightly higher in the statin-exposed group (24.9 [SD 0.4]) compared to in the non-exposed group (23.9 [SD 0.3]). There was a similar number of smokers and less than five women with diabetes prior to pregnancy in both groups (Table 3).

There was no difference in the risk of preeclampsia, gestational hypertension, gestational diabetes, or intrapartum hemorrhage between pregnancies exposed-and not exposed to statins. Low birth weight occurred more frequently in pregnancies exposed to statins (OR 2.20 [95% CI: 1.11, 4.49]). There were similarly, although non-significant, higher proportion of the statin exposed pregnancies that experienced preterm birth (OR 1.74 [95% CI: 0.93, 3.24]) and proportion with birth weight below the 10^th^ percentile (OR 1.33 [95% CI: 0.78, 2.20]).

Among the statin exposed pregnancies, 44% (n= 8) of the premature births had c-section as delivery method, whereas the corresponding numbers in the non-exposed group was 15% (n = 37). Thus, c-section was more common among the statin-exposed pregnancies (OR was 1.55 [95% CI: 1.03, 2.31]). The results for subgroups of c-section were however divergent: there was a non-significant higher OR for acute c-section (OR 1.25 [95%CI: 0.6, 2.5]), and non-significant lower odds for scheduled/planned c-section (0.7 [95%CI: 0.4, 1.6]) among the statin exposed pregnancies.

The number of cases was too low to investigate the risk of neonatal deaths, stillbirths and congenital heart defects in statin exposed versus non-exposed pregnancies among women with FH.

### Statin use in controls

Statins were used in 16 pregnancies among controls, of whom less than five had a low birthweight offspring and/or delivered a pre-term baby. Due to the small sample size, no further statistical analysis of associations with statin exposure among controls was performed.

## Discussion

With data on 3869 pregnancies among women with genetically verified FH, this is the largest study to date on pregnancy outcomes in women with FH. FH was associated with lower risk of gestational diabetes and intrapartum hemorrhage during birth and higher, non-significantly risk of preeclampsia compared to controls. There was no excess risk for other adverse pregnancy-related outcomes in either the mothers with FH, or their offspring. Statin use during pregnancy was associated with a higher risk of having offspring with low birth weight, as well as a (non-significantly) higher proportion of preterm births, compared to non-exposed pregnancies.

Previous studies showing an association between hypercholesterolemia and preeclampsia and gestational diabetes in general populations ^8, 9, 27^ may be limited by reverse causality and confounding factors because hypercholesterolemia is strongly associated with an unhealthy dietary pattern, age, physical inactivity, insulin resistance and overweight and obesity – all well-known risk factors for preeclampsia ^11, 28^ and gestational diabetes ^10, 29^. The genetic predisposition that leads to FH allows us to study the role of LDL-C in risk of adverse pregnancy outcomes less biased of reverse causality. Moreover, in the current study, the common risk factor BMI at start and end of pregnancy were similar among the FH and control populations, and there was a very low percentage with diabetes mellitus before pregnancy in both groups. Furthermore, there was no significant interaction between estimated known FH status and risk of any outcomes, ruling out possible detection bias as women with FH may be more closely followed-up in the health-care system in general.

Nonetheless, (chronic) hypertension, the main risk factor for preeclampsia ^28^ was somewhat higher among women with FH (0.5%) compared to controls (0.3%), despite that similar frequencies of hypertension (not limited to chronic) in the total FH population compared to controls have previously been reported ^30^.

In 2011 we reported that women with FH had no excess risk of gestational diabetes during follow-up (1967-2006). ^12^. However, diagnostics and registration of gestational diabetes in Norway have improved after 2006, and the prevalence has significantly and simultaneously increased annually according to Medical Birth Registry statistics^31^. As an example, between 2010 and 2016 the prevalence of gestational diabetes increased from 2.6% to 6%^32^. Given that we in the present study report risk of gestational diabetes between 1967 and 2018, the association between FH and gestational diabetes may be underestimated.

The observed 19% lower risk of intrapartum hemorrhage during delivery in the FH population was somewhat surprising. The difference cannot be explained by some major risk factors (multiple pregnancies, older age, high BMI, IVF, high birth weight babies, c-section, preeclampsia) ^33^ as the occurrence of these risk factors were similar among FH and controls. In search of an explanation, we looked to the well-established association between high levels of Lp(a) and lower risk of significant hemorrhage in general ^34, 35^. This could be relevant as high Lp(a) levels has been observed in the Norwegian FH cohort ^36–38^. However, high Lp(a) is not a general trait in all individuals with genetically verified FH ^39, 40^.

The observed borderline higher risk of preeclampsia in women with FH, extending our previous non-significant finding in a smaller sample of the population group ^12^, and the novel findings of lower risk of gestational diabetes and intrapartum hemorrhage, may therefore represent an argument that high LDL-C in itself is a player and not only a bystander in the development of these pregnancy-related disorders.

### Adverse outcomes in offspring

In line with our previous work ^12^, and compared to a matched control group, we found that women with genetically verified FH did not have a significantly higher risk of low birth weight or preterm delivery adjusting for the mothers’ age at pregnancy, parity and children’s birth year. The FH population did not have a higher risk of having infants with congenital (or any) malformations compared to controls, although clear conclusions cannot be drawn due to low number of cases.

We observed a 2.2 (95% CI: 1.11, 4.49) higher odds of low birth weight in offspring from mothers with FH with statin prescriptions dispensed during pregnancy, compared to the offspring of the non-exposed mothers. In relation to this, a higher odd of c-section (OR 1.55 (95% CI: 1.03, 2.31)) and a non-significantly higher odds of preterm birth (OR 1.74 (95% CI: 0.93, 3.24)) were also associated with statin exposure in pregnancy. The associations between statin use in pregnancy and low birth weight and preterm birth are supported by similar findings in 469 statin exposed women in Taiwan ^22^, and a recent a real-world pharmacovigilance study based on the Food and Drug Administration adverse effects reporting system ^21^. On the contrary, a meta-analysis showed that statin use was associated with reduced (although non-significantly) risk of preterm delivery ^41^. One previous study has investigated adverse outcomes associations with statin use in women with FH: In 39 pregnancies, no adverse pregnancy outcomes was associated with statin use ^6^, although statistical power is a limitation here. It has been suggested that the physiological state of FH could impact the risk of premature births in relation to placental insufficiency ^42^, however as we only observed excess risk among the statin-exposed pregnancies in women with FH (and not in the total FH population) the underlying cause of this association should be further investigated.

Unhealthy lifestyle behaviors such as smoking ^43^ in addition to high BMI and/or excess weight gain during pregnancy, and babies born to mothers with diabetes are often larger than expected for the gestational age, are all well known risk factors for low birth weight ^44^. Except for slightly higher pre-pregnancy BMI among the statin-exposed pregnancies, all other risk factors were similar in the statin-exposed and non-exposed group. Still, we cannot rule out that there are other possible explanations for these associations. Almost all women in our sample used statins due to their FH diagnosis, but since statins are generally not recommended during pregnancy, those who continued statins during pregnancy may differ in health behavior from their non-exposed counterparts.

### Strengths and limitations

A major strength of the study is the use of MBRN registry data dated back to 1967 for all pregnancies (from week 12-16 [depending on year of pregnancy]) in large cohorts of women with and without FH residing in Norway. The MBRN contains a large amount of information on pregnancy-related outcomes both in the mother and the offspring ^25^. We also obtained data from the Norwegian patient registry on occurrence of coronary heart disease, stroke, pulmonary embolism, deep vein thrombosis in association with pregnancy, however there were too few cases to be included in the current study. Since statin use often is self-reported and hence underestimated ^12^ and statin use during pregnancy in most cases probably is unintentional, it is a major strength that we have objective data on pharmacy-dispensed statins. Almost all adult women with a molecular genetic FH diagnosis are recommended to use statins. Thus, use of statins due to other comorbidities like diabetes play a minor role. However, we do not know for certain that the women took their statins as prescribed.

Since the registration period for variables in the MBRN varies (e.g. we only have smoking information after 1999), we do not have complete datasets on all relevant exposure measures. Similarly, there were too few cases of diabetes mellitus (all types) and chronic hypertension prior to pregnancy to be able to adjust for this in the analysis. As we do not have complete coverage of all comorbidities (including diabetes and hypertension diagnosis from other sources than the MBRN), we cannot rule out that some of the associations observed can be driven by pre-existing metabolic diseases. Since we do not have information on all lifestyle habits, we cannot rule out residual confounding for our reported associations. This may be particularly important for the associations between statin use and adverse pregnancy outcomes.

To sum up, this longitudinal registry-based study demonstrates that women with genetically verified FH have lower risk of gestational diabetes and intrapartum hemorrhage during birth, but may have elevated risk of preeclampsia compared to controls, suggesting a potential role of high LDL as a common risk factor. Reassuringly, no excess risk of adverse pregnancy outcomes in offspring from mothers with FH were observed. However, statin use in pregnancy among women with FH was associated with a higher risk of having offspring with low birthweight. Although these results can be subject to confounding by lifestyle factors, it may still support a continued precautionary approach towards statin use when planning and during pregnancy. The higher frequency of preterm births among the statin exposed pregnancies may be an explanation for the observed association that warrants further investigation.

## Acknowledgments

Data is obtained from several Norwegian health registries. However, the registries have no role in the interpretation and presentation of data.

## Data availability statement

The anonymous data underlying this article may be obtained upon reasonable request to the corresponding author.

---

**Figure 1.** Flowchart of study inclusion of women with familial hypercholesterolemia (FH) and age-matched controls.

**Figure 2.** Adverse pregnancy outcomes in women with familial hypercholesterolemia (FH) versus controls. OR= odds ratio retrieved from logistic regression with clustered robust standard error. Adjusted model= adjusted for mothers’ age at pregnancy and children’s birth year in categories and parity. Large bleedings/ intrapartum hemorrhage = (>500 ml) during delivery.

**Figure 3.** Adverse pregnancy outcomes in offspring from mothers with familial hypercholesterolemia (FH) versus controls. OR= odds ratio retrieved from logistic regression with clustered robust standard error. Adjusted model= adjusted for mothers age at pregnancy and children’s birth year in categories and parity. Preterm birth =Birth in gestational week 37 or less. Any malformations = minor and major malformations combined.

**Figure 4.** Number of prescriptions fills per 2-week period from one year before conception to one year after birth in the FH population (n=1051). Conception is set to time zero, and the bars are colored by prescription fill inside (red) or outside (blue) pregnancy, or those prescription fill periods that overlap (carry-over) up to 42 days into pregnancy (green).

**Figure 5.** Risk of adverse pregnancy outcomes in pregnancies exposed to statins* (n=260) versus pregnancies not exposed to statins (n=791) among women with FH. Odds ratio (OR) retrieved from logistic regression, clustered robust standard error. Statin exposure: filling at least one prescription for statins either during pregnancy, or before conception if the prescription package duration would last into pregnancy. Adjusted model = adjusted for mothers age at pregnancy and children’s birth year in categories and parity. Large bleedings/ intrapartum hemorrhage = >500 ml during delivery. Preterm birth = birth in ≤gestational week 37.

